# Optimal piecewise constant vaccination and lockdown policies for COVID-19

**DOI:** 10.1101/2021.01.13.21249773

**Authors:** Gabriel A. Salcedo-Varela, F. Peñuñuri, D. González-Sánchez, Saúl Díaz-Infante

**Affiliations:** Departamento de Matemáticas, Universidad de Sonora, Blvd. Luis encinas y Rosales S/N, Hermosillo, Sonora, México, C.P. 83000; Facultad de Ingeniería, Universidad Autónoma de Yucatán, A.P. 150, Cordemex, Mérida, Yucatán, México; CONACYT-Universidad de Sonora, Departamento de Matemáticas, Blvd. Luis encinas y Rosales S/N, Hermosillo, Sonora, México, C.P. 83000

**Keywords:** COVID-19, Optimal Control, COVAX, Vaccination, WHO-SAGE, DALYs

## Abstract

We formulate a controlled system of ordinary differential equations, with vaccination and lockdown interventions as controls, to simulate the mitigation of COVID-19. The performance of the controls is measured through a cost functional involving vaccination and lockdown costs as well as the burden of COVID19 quantified in DALYs. We calibrate parameters with data from Mexico City and Valle de Mexico. By using differential evolution, we minimize the cost functional subject to the controlled system and find optimal policies that are constant in time intervals of a given size. The main advantage of these policies relies on its practical implementation since the health authority has to make only a finite number of different decisions. Our methodology to find optimal policies is relatively general, allowing changes in the dynamics, the cost functional, or the frequency the policymaker changes actions.

## 1. Introduction

At the date of writing this manuscript, the USA is running its COVID-19 Vaccination with Pfizer-BioNTech vaccine. This vaccine development, along with Astra-Zeneca, Cansino, Sputnik V, and Novavax, promises to deliver enough doses for Latin America. In Mexico, particularly, the first stock with around 40 000 shots has arrived past Christmas. In past October, WHO established a recommended protocol for prioritizing access to this pharmaceutical hope, giving clear lines about who has to be vaccinated first and why. However, each developed vaccine implies different issues around its application. For example, the Pfizer-BioNTech vaccine requires two doses and particular logistics requirements that demand special services. In Mexico, despite Pfizer-BioNTech, has been taking the responsibility to capacitate personnel that manages the Vaccination, there is an explicit demand for logistics resources that limit the institutions’ response. On the other hand, nonpharmaceutical interventions (NPIs), like Lockdown, mobility reduction, social distancing, and other restrictions, also involve economic costs. Our research in this manuscript explores the effect of two interventions, Vaccination and Lockdown, to mitigate the propagation of COVID-19.

Among the related literature about the two interventions we deal with in this paper, we can mention the following. The problem of who is vaccinated first, when the number of available shots is limited, has been transformed into an optimal allocation problem of vaccine doses in [1–3]. These articles face the critical question: how much doses allocate to each different group according to risk and age to minimize the burden of COVID-19. Our study takes the allocation for granted and modulates vaccination and lockdown-release rates as a combined strategy. Further, papers modeling NPIs consider the diminish of contact rates by reducing mobility or modulating parameters regarding the generation of new infections by linear controls [4, 5], lockdown–quarantine [6], and shield immunity [7]. Also, Libotte et al. report in [8] optimal vaccination strategies for COVID-19.

Since health services’ response will be limited by the vaccine stock and logistics costs, implementing in parallel NPIs is imminent. We focus on formulating and studying via simulation a lockdown-vaccination system by consider the vaccine recently approved by Mexico Health Council. We aim to design a dose application schedule subject to a given vaccine stock applied in a given period. For this purpose, we formulate an optimal control problem that minimizes the burden of COVID-19 in DALYs [9], the cost generated by running the vaccination campaign, and economic damages due to lockdown.

One of the main features of our model is that we consider piecewise constant control policies instead of general measurable control policies—also called permanent controls—to minimize a cost functional. General control policies are difficult to implement since the authority has to make different choices every permanently. The optimal policies we find are constant in each interval of time and hence these policies are easier to implement. To the best of our knowledge, our manuscript is the first optimal control model with both lockdown and vaccination strategies.

In Section 2, we formulate the basic spread model for COVID19 and calibrate its parameters. Then, Section 3 establishes the lockdown–vaccination model and discusses the regarding reproductive number in Section 4. In Section 5 we describe the optimal control problem which consists in minimizing a cost functional subject to controlled lockdown–vaccination system. The optimal policies we find, by solving numerically the optimal control problem, are presented in Section 6. We conclude with some final comments in Section 7.

## 2. Covid-19 spread dynamics

We split a given population of size *N* in the basic SEIR structure with segregated classes according to the manifestation of symptoms. Let *L, S, E, I*_*S*_, *I*_*A*_, *H, R, D* respectively denote the class of individuals according to their current state, namely

**Lockdown** (*L*) All individuals that have low or null mobility and remain under isolation. Thus individuals in this class reduce their contagion probability.

**Suceptible** (*S*) Individuals under risk

**Exposed** (*E*) Population fraction that hosts SARS-CoV-2 but cannot infect

**Infected-Symptomatic** (*I*_*S*_) Population infected fraction with symptoms and reported as confirmed cases

**Infected-Asymptomatic** (*I*_*A*_) Infected individuals with transitory or null symptoms and unreported

**Hospitalized** (*H*) Infected population that requires hospitalization or intensive care.

**Recover or removed** (*R*) Population that recovers from infection and develops partial immunity

**Death** (*D*) Population fraction that died due to COVID-19

To fit data of cumulative reported symptomatic cases, we postulate the counter state 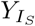 and make the following assumptions.

### Assumptions 1.

According to above compartment description, we made the following hypotheses.

(A-1) We suppose that at least 30 % of the population is locked down and a fraction of this class eventually moves to the susceptible compartment at rate *δ*_*L*_.

(A-2) Force infection is defined as the probability of acquiring COVID-19 given the contact with a symptomatic or asymptomatic individual. Thus we normalize with respect to alive population population *N*

(A-3) Susceptible individuals become exposed—but not infectious—when they are in contact with asymptomatic or symptomatic individuals. Thus *β*_*S*_ and *β*_*A*_ denote the probabilities of being infectious given the contact with a symptomatic or asymptomatic infectious individuals, respectively.

(A-4) After a period of latency 1*/κ* = 5.1 days, an exposed individual becomes infected. Being *p* the probability of developing symptoms and (1 − *p*) the probability of becoming infectious but asymptomatic. Thus *pκe* denotes the exposed individuals that become infectious and develop symptoms.

(A-5) Asymptomatic individuals do not die or stay in the hospital.

(A-6) A fraction *µ*_*H*_ of symptomatic individuals dies due to COVID-19 without hospitalization.

Thus we formulate the following Ordinary Differential Equation (ODE), see Figure 1, to complete the underlying description.

**Figure 1:**
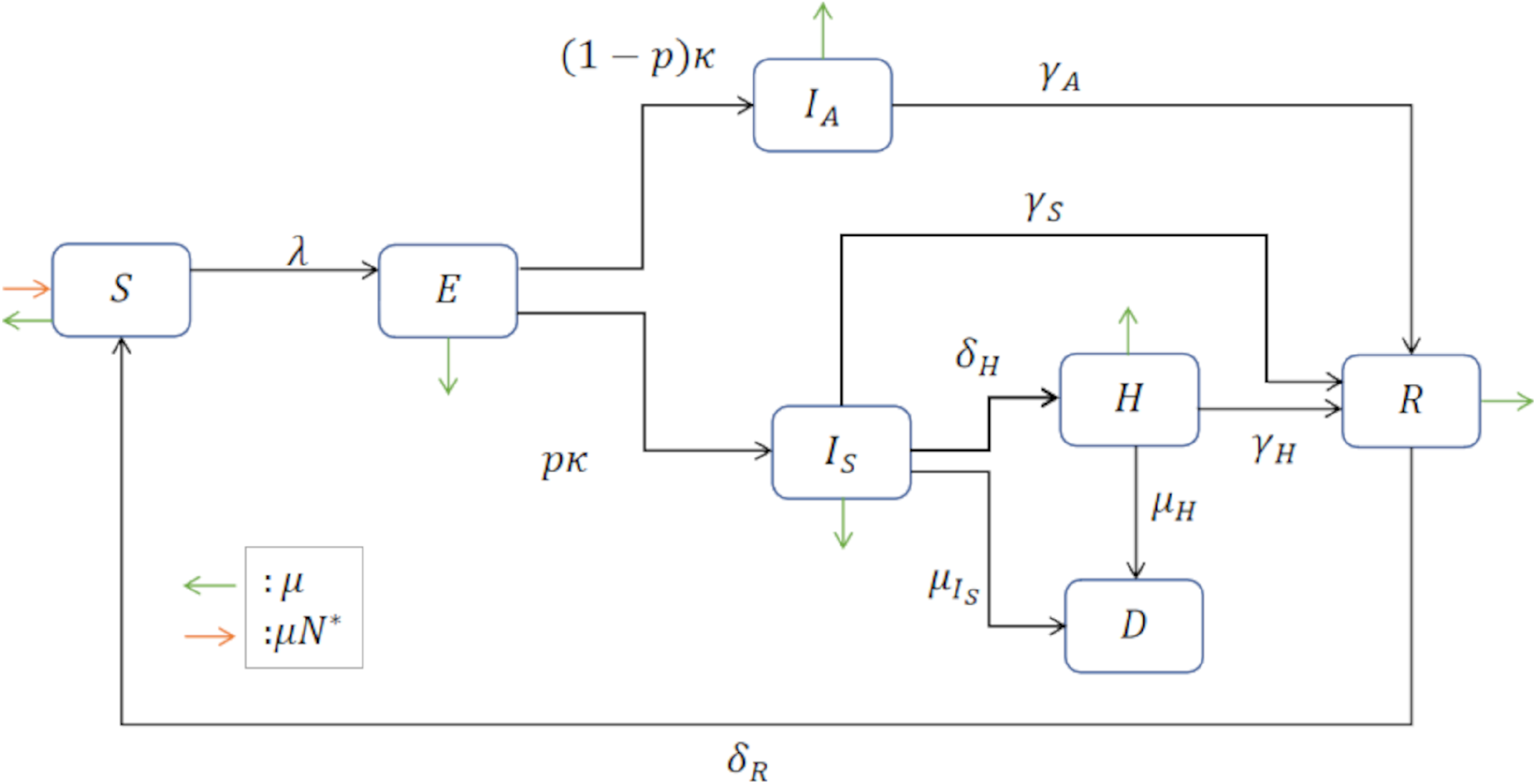
Compartmental diagram of COVID-19 transmission dynamics. Consider the class: Susceptible (*S*), exposed (*E*), symptomatic infected (*IS*), asymptomatic infected (*IA*), recovered (*ℝ*), death (*D*) and vaccinated (*V*) individuals. It is important to mention that *IS* represents the proportion of symptomatic individuals who will later report to some health medical center.

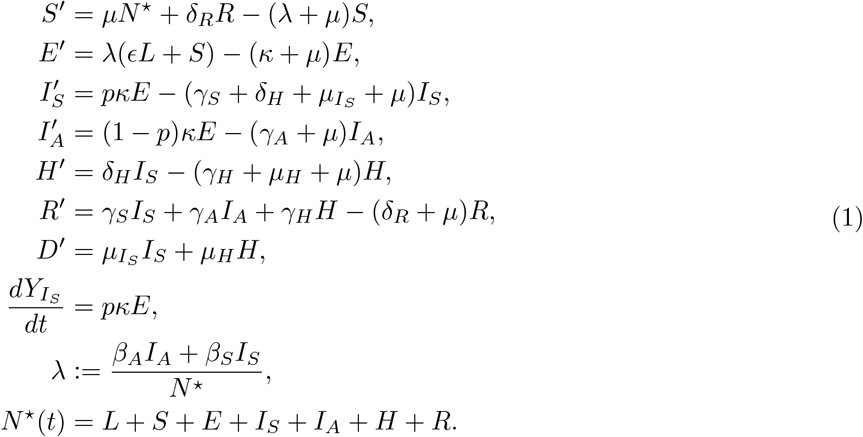

We display in Figure 2 a typical evolution of this COVID-19 spreed dynamics. Table 1 encloses notation and reference values.

**Table 1:** Parameters definition of model in Equation (1).

| Parameter | Description |
| --- | --- |
| $\mu$ | Death rate |
| $\beta_S$ | Infection rate between susceptible and symptomatic infected |
| $\beta_A$ | Infection rate between susceptible and asymptomatic infected |
| $\lambda_V$ | Vaccination rate |
| $\delta_V^{-1}$ | Vaccine-induced immunity |
| $\varepsilon$ | Vaccine efficacy |
| $\kappa^{-1}$ | Average incubation time |
| $p$ | New asymptomatic generation proportion |
| $\theta$ | Proportion of susceptible individuals under lockdown |
| $\gamma_S^{-1}$ | Average time of symptomatic recovery |
| $\gamma_A^{-1}$ | Recovery average time of asymptomatic individuals |
| $\gamma_H^{-1}$ | Recovery average time by hospitalization |
| $\delta_R^{-1}$ | Natural immunity |
| $\delta_H$ | Infected symptomatic hospitalization rate |

**Figure 2:**
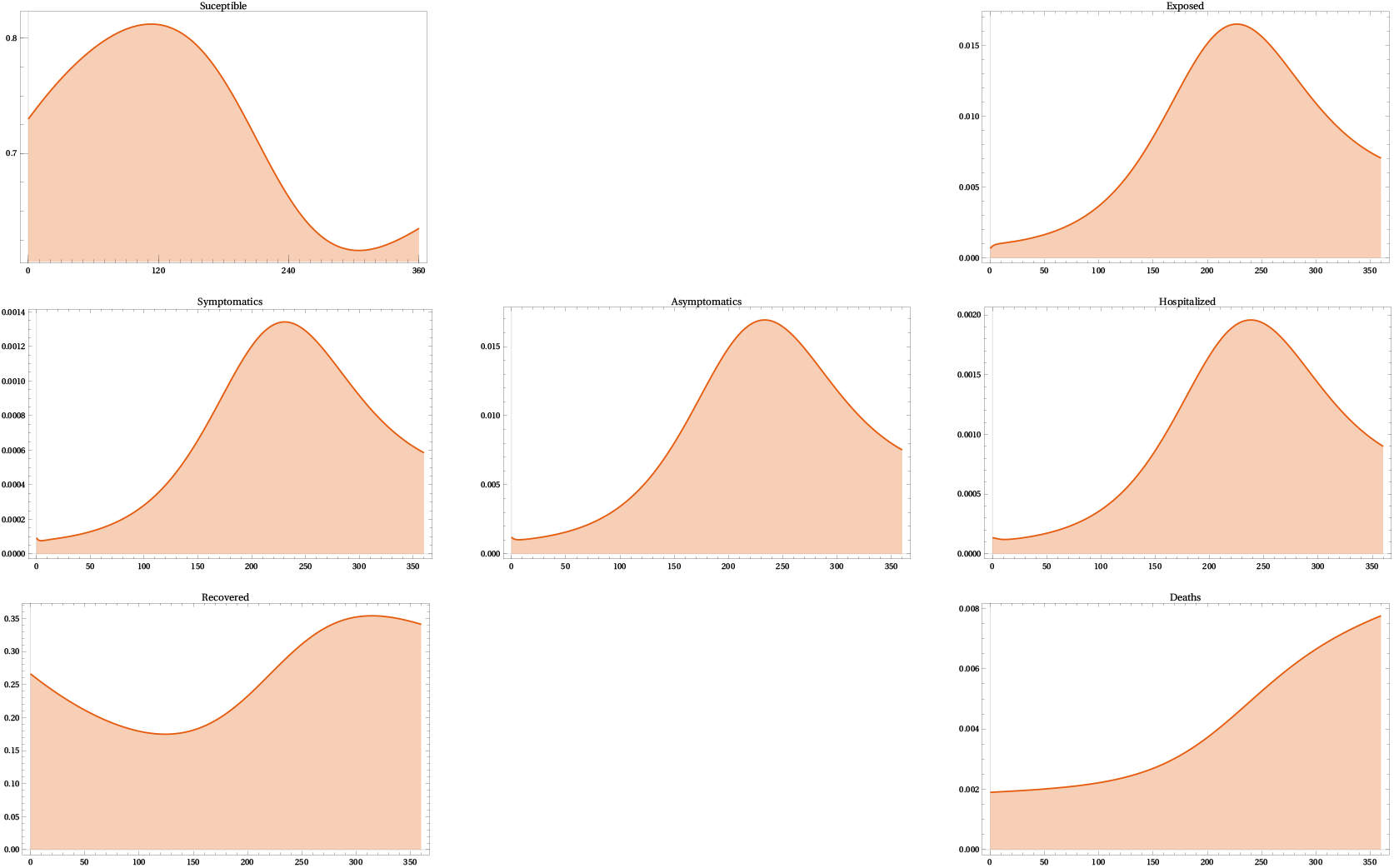
Spreed dynamics of COVID-19 according to model in Equation (1).

### 2.1. Parameter calibration

We calibrate parameters of our base dynamics (1) via Multichain Montecarlo (MCMC). To this end, we assume that the cumulative incidence of new infected symptomatic cases *CI*_*S*_ follows a Poisson distribution with mean *λ*_*t*_ = *IC*_*s*_(*t*). Further, following ideas from [11] we postulate priors for *p* and *κ* and count the cumulative reported-confirmed cases in the CDMX-Valle de Mexico database [10]

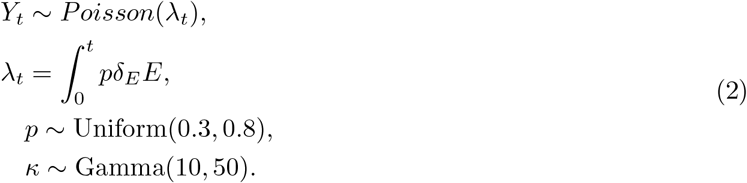

Using Van den Driessche’s [12] definition of reproductive number we obtain

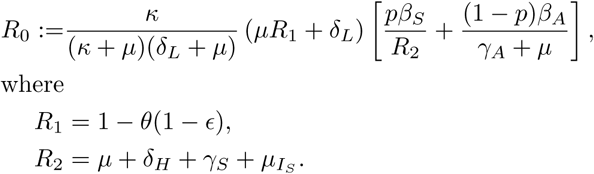

Figure 3 displays data of cumulative confirmed cases of COVID-19 in Mexico city, and Figure 4 displays the fitted curve of our model in Equations (1) and (2). Table 2 encloses estimated parameters to this setting.

**Table 2:**
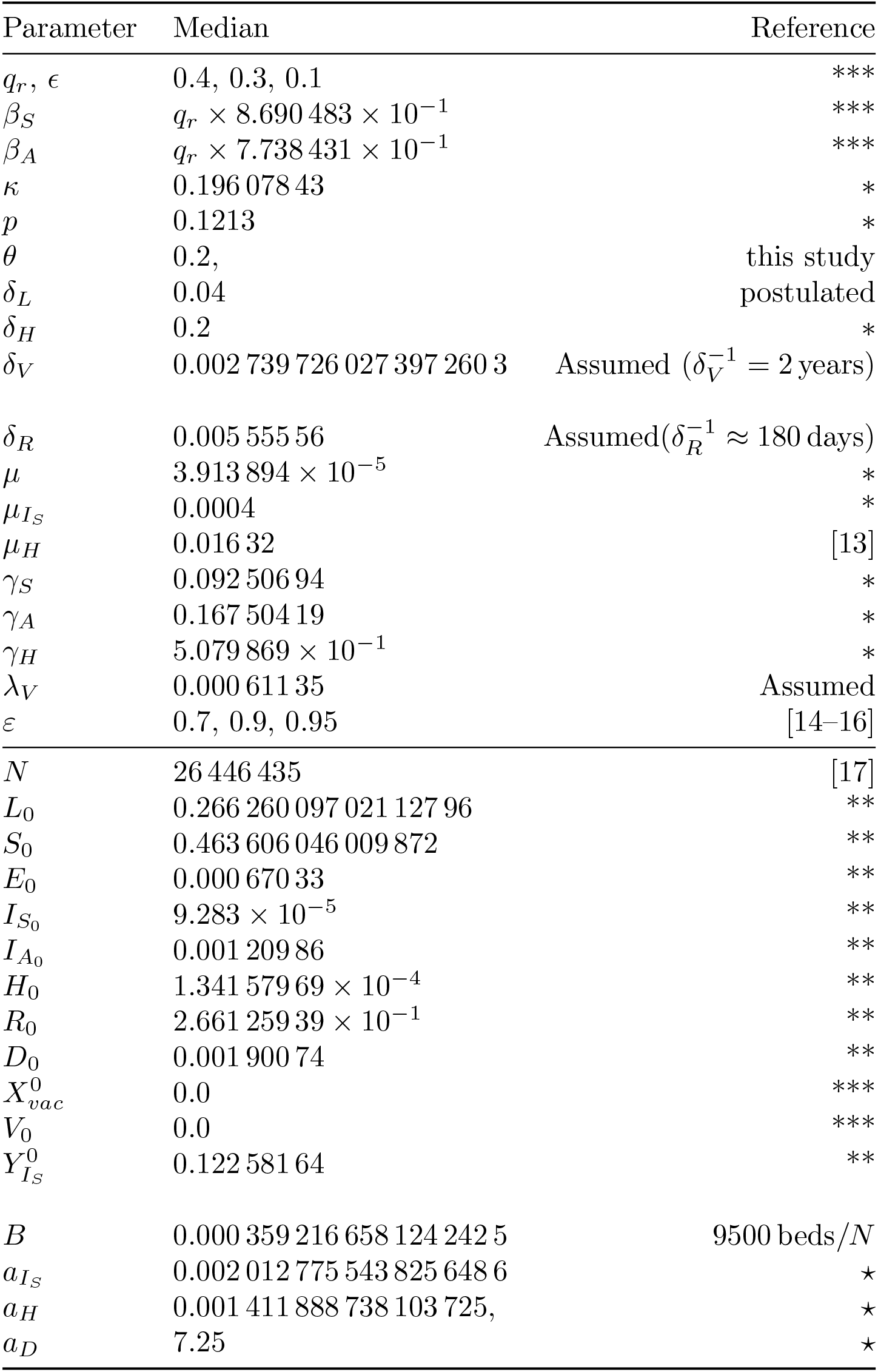
Model parameters. (*) Values based mainly in [13, 18]. (**) Estimated. (***) This study. (⋆) From [19].

**Figure 3:**
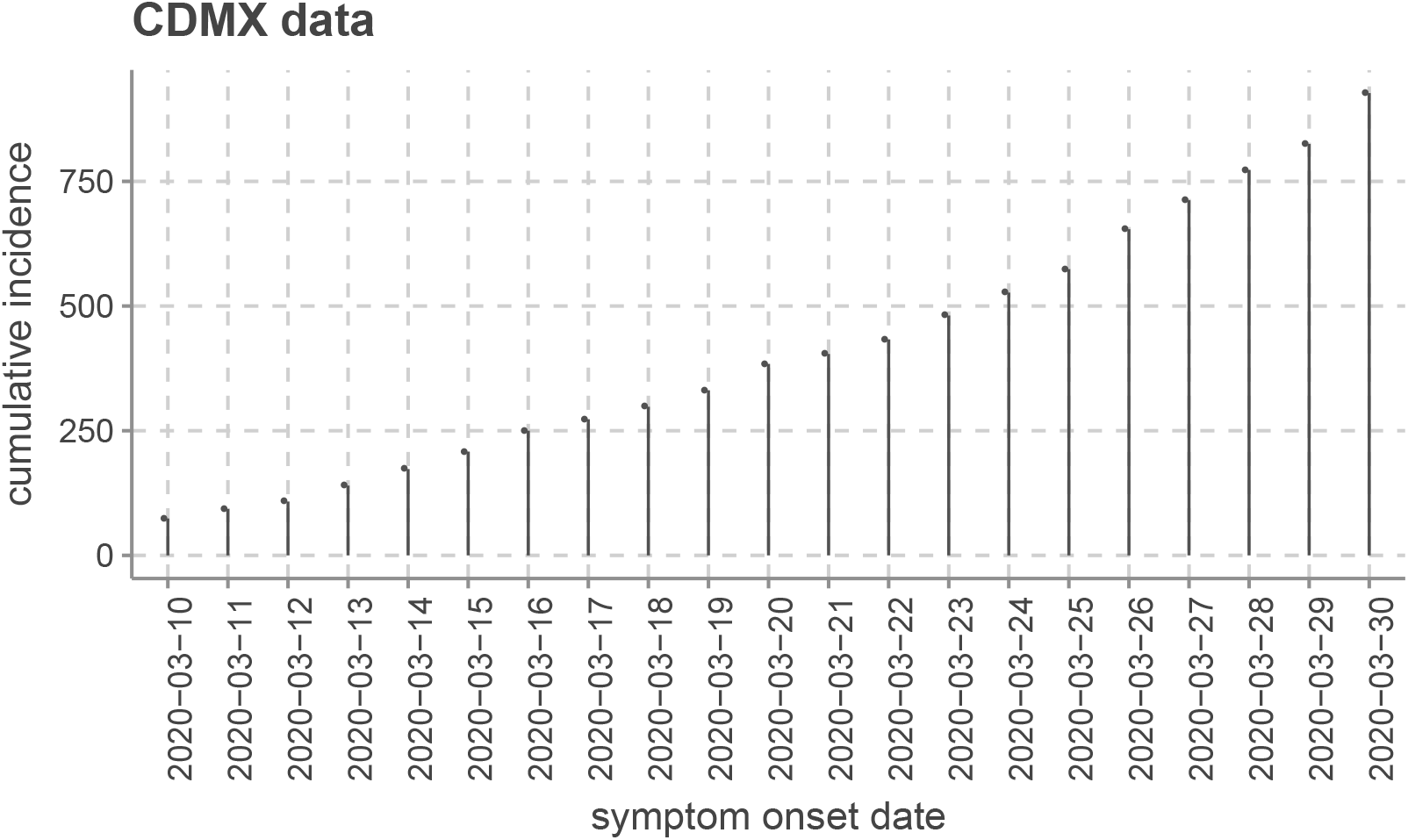
Cumulative new symptomatic and confirmed COVID19 reported cases from Ciudad de Mexico and Valle de Mexico [10] between March, 10, to March 30 of 2020. https://plotly.com/AdrianSalcedo/48/

**Figure 4:**
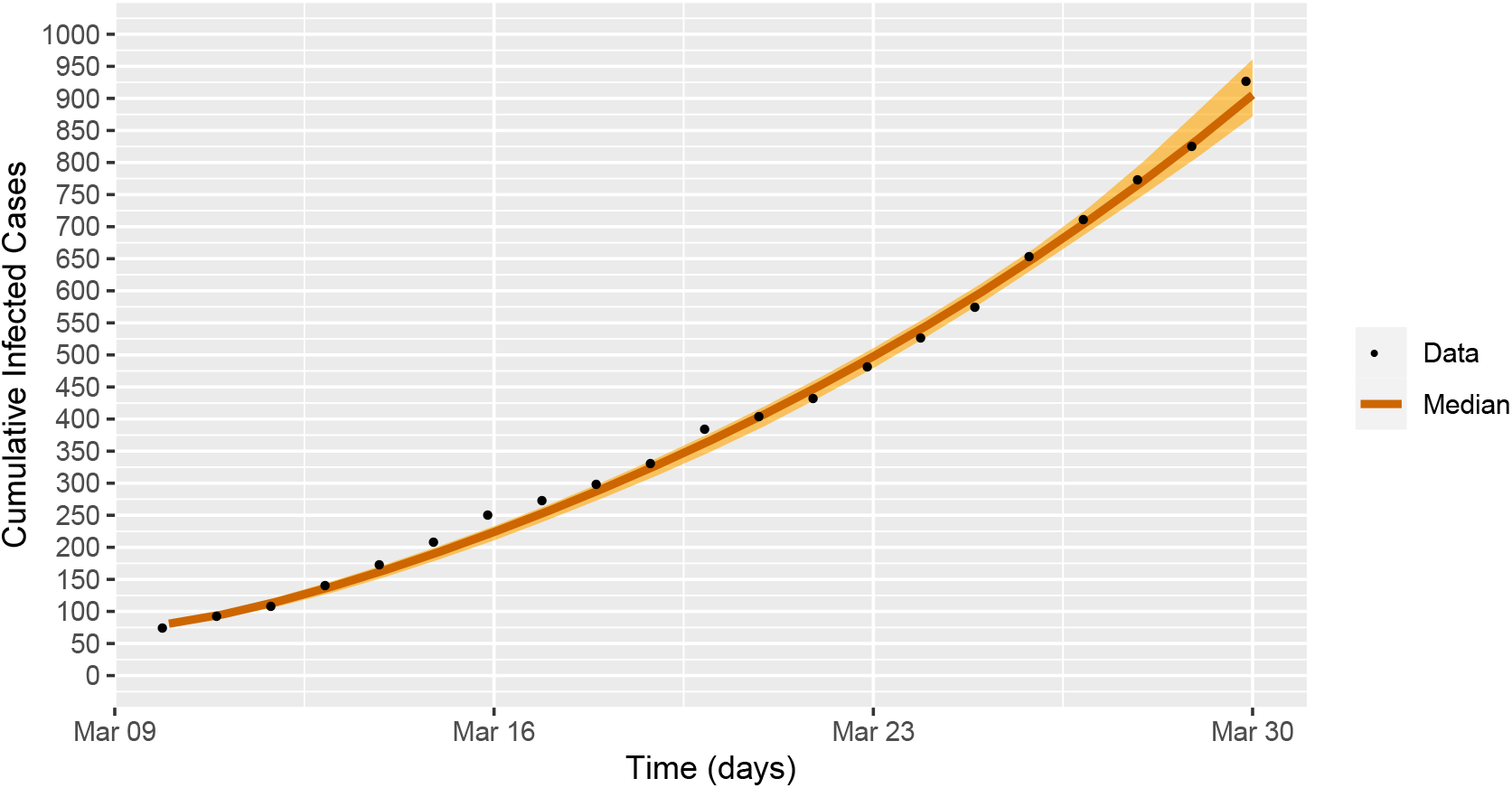
Fit of daily new cases of Mexico city during exponential growth.

## 3. Imperfect-preventive COVID-19 vaccination

### Assumptions 2.

According to COVID-19 dynamics Equation (1), we made the following modeling hypotheses about the regarding vaccine.

(VH-1) Vaccine is preventive and only reduce susceptibility.

(VH-2) The vaccination camping omits testing to detect seroprevalence. Thus Exposed, Infected Asymptomatic and Recovered Asymptomatic individuals are undetected but would obtain a vaccine dose —which in this model represents a waste of resources

(VH-3) Individuals under lockdown are also vaccinated

(VH-4) The vaccine is leaky and with efficacy *ϵ* ∈ [0.7, .975]

(VH-5) Vaccine induced immunity last 2 years

(VH-6) Natural immunity last a period of 180 days

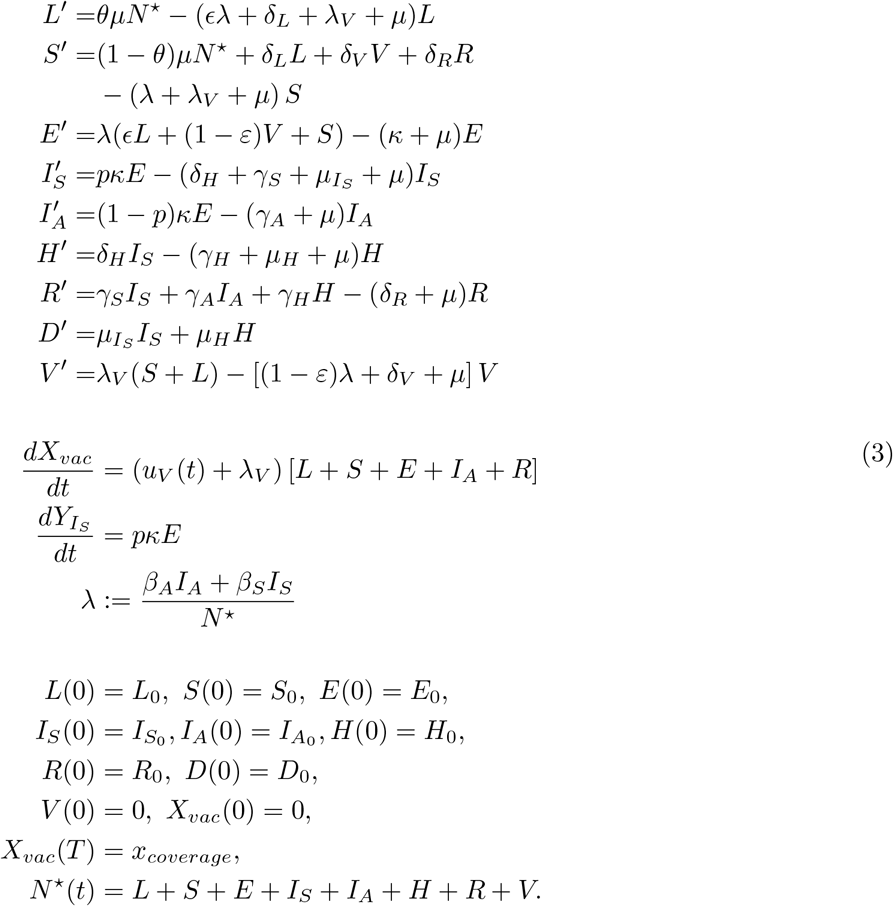

## 4. Lockdown-Vaccination reproductive number

The basic reproductive number, which is generally denoted by *R*_0_, is a threshold quantity we can control with particular strategies. The epidemiological interpretation of *R*_0_ is the average number of secondary cases produced by an infected individual introduced into a population of susceptible individuals. Using van den Driessche’s [20] definition of reproductive number we obtain

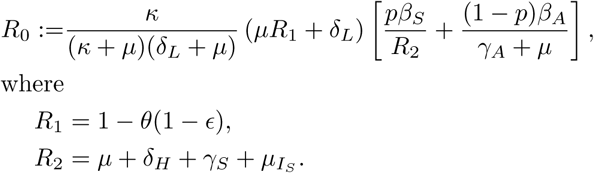

The factor 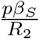 measures the proportion of new infections generated by a symptomatic infectious individual in the time that it lasts infected. In a similar way, the factor 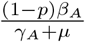 measures the new infections generated by an asymptomatic infectious individual in the time that it lasts infected. The factor 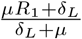 measures the number of individuals in lockdown that leave the lockdown, which can be infected. And finally, the factor 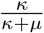 measures the time of the disease’s incubation. If we consider that there is no lockdown, then *R*_0_ is reduced to

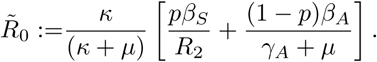

Note that we have the relation 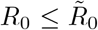. These indicate that there is greater transmission of the disease if there is no lockdown.

Considering Assumptions 2, we can establish a vaccine reproductive number, in which individuals who have already been vaccinated can become infected individuals by being in contact with the symptomatic infected. Using van den Driessche’s [20] definition of reproductive number and [21], we obtain

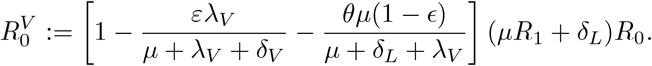

The threshold quantity 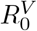is the reproductive number of infection which can be interpreted as the number of infected people produced by one infected individual introduced into the population in the presence of vaccination.

Figure 6 shows 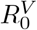as a function of vaccine efficacy and vaccination rate. The blue line, *ε* = 0.8, tells uswhat value of *λ*_*V*_ we take to have the level curve where 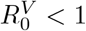.

**Figure 5:**
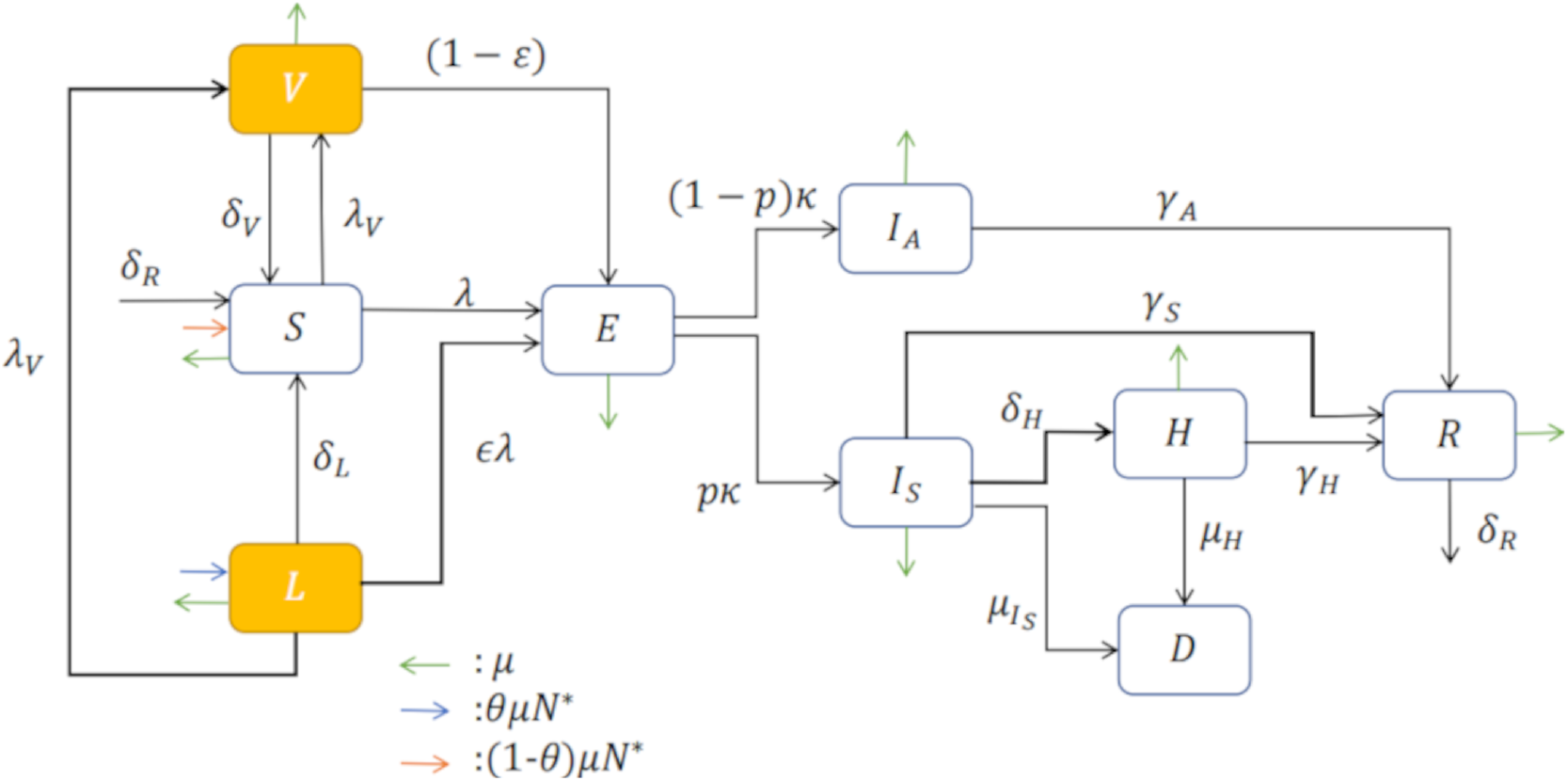
Compartmental diagram of COVID-19 transmission dynamics which including vaccination dynamics. Here, we consider the Lockdown class (*L*).

**Figure 6:**
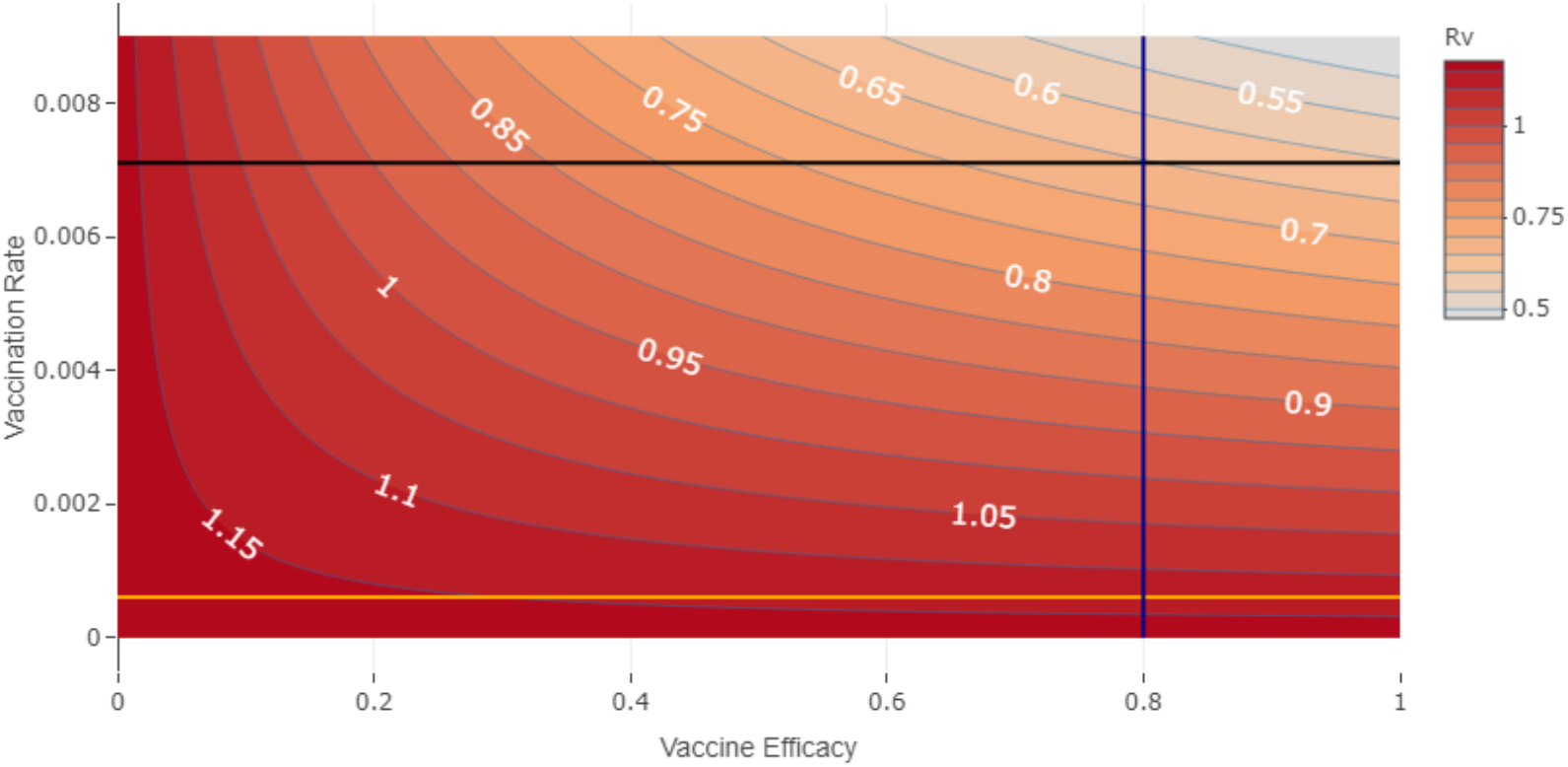
Contour plot of 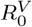 as a function of vaccine efficacy (*ε*) and vaccination rate (*λ*_*V*_) and vaccine-induced immunity average time of half year. Orange line represents the value of *λ*_*V base*_ = 0.000 611, corresponding to a coverage *x*_*coverage*_ = 0.2 and a horizon time *T* = 365 days. Intersection of black line and blue line show a scenario in which it is possible to have the 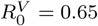, considering a vaccine efficacy of 0.8 and a vaccination rate of 0.7.

Figure 7 shows the region of vaccine efficacy and vaccination rate, for which 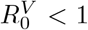. In this scenario, there is no lockdown effect. Note that when vaccine efficacy is less than 0.2, there is no vaccination rate for which 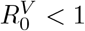. In the absence of lockdown individuals, there is a region not feasible for vaccine efficacy and vaccination rate.

**Figure 7:**
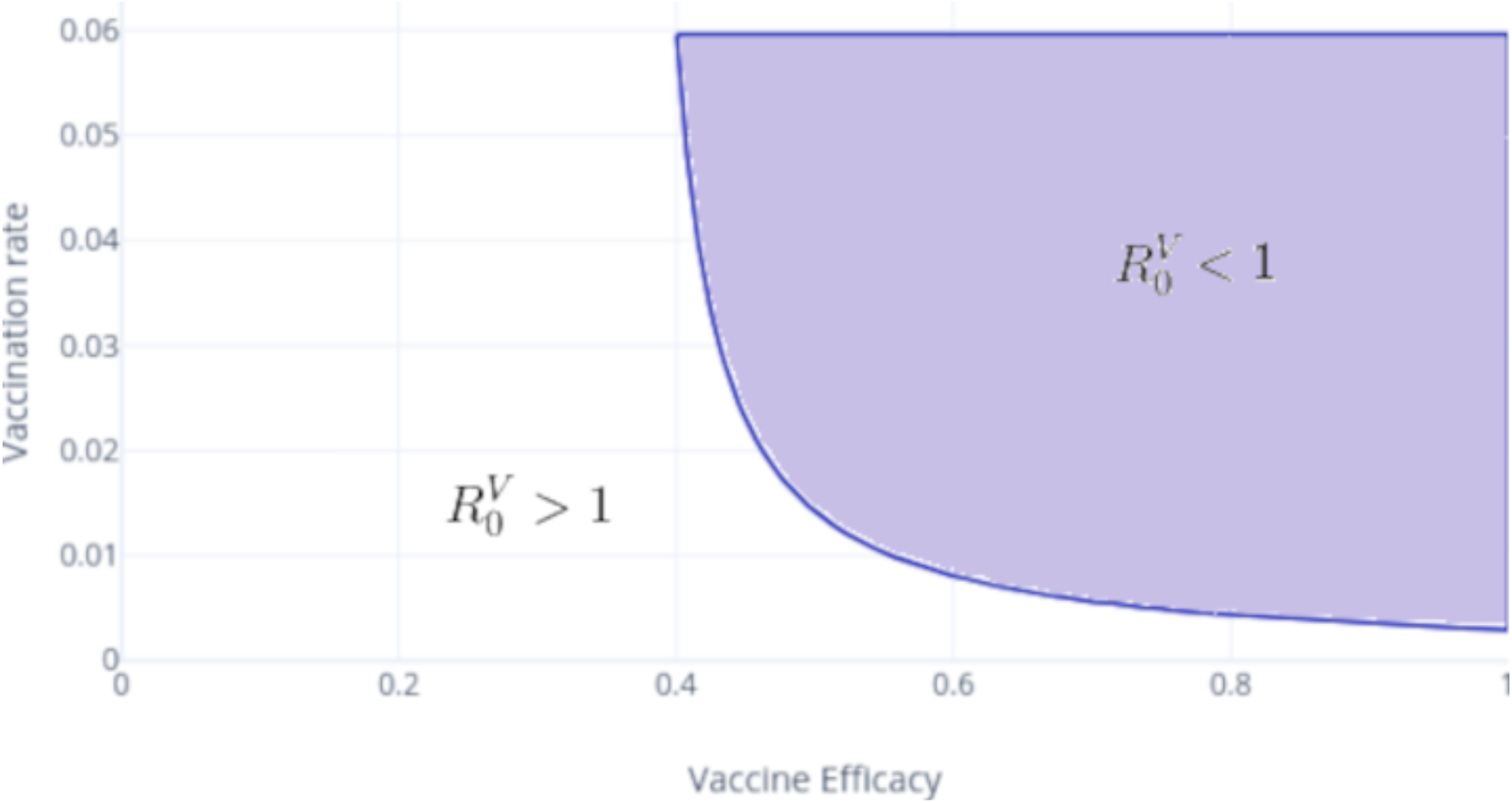
Vaccine efficacy versus vaccination rate feasibility. In the purple shaded region 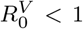 and in the white region 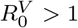. Note that, for this scenario, we consider no lockdown individuals. https://plotly.com/AdrianSalcedo/85/

Figure 8 shows the region for which 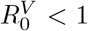 when there are lockdown individuals. In this scenario, weobserve that there are values for the vaccine efficacy and vaccination rate where 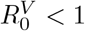, which are smallerthan the ones we choose in Figure 7.

**Figure 8:**
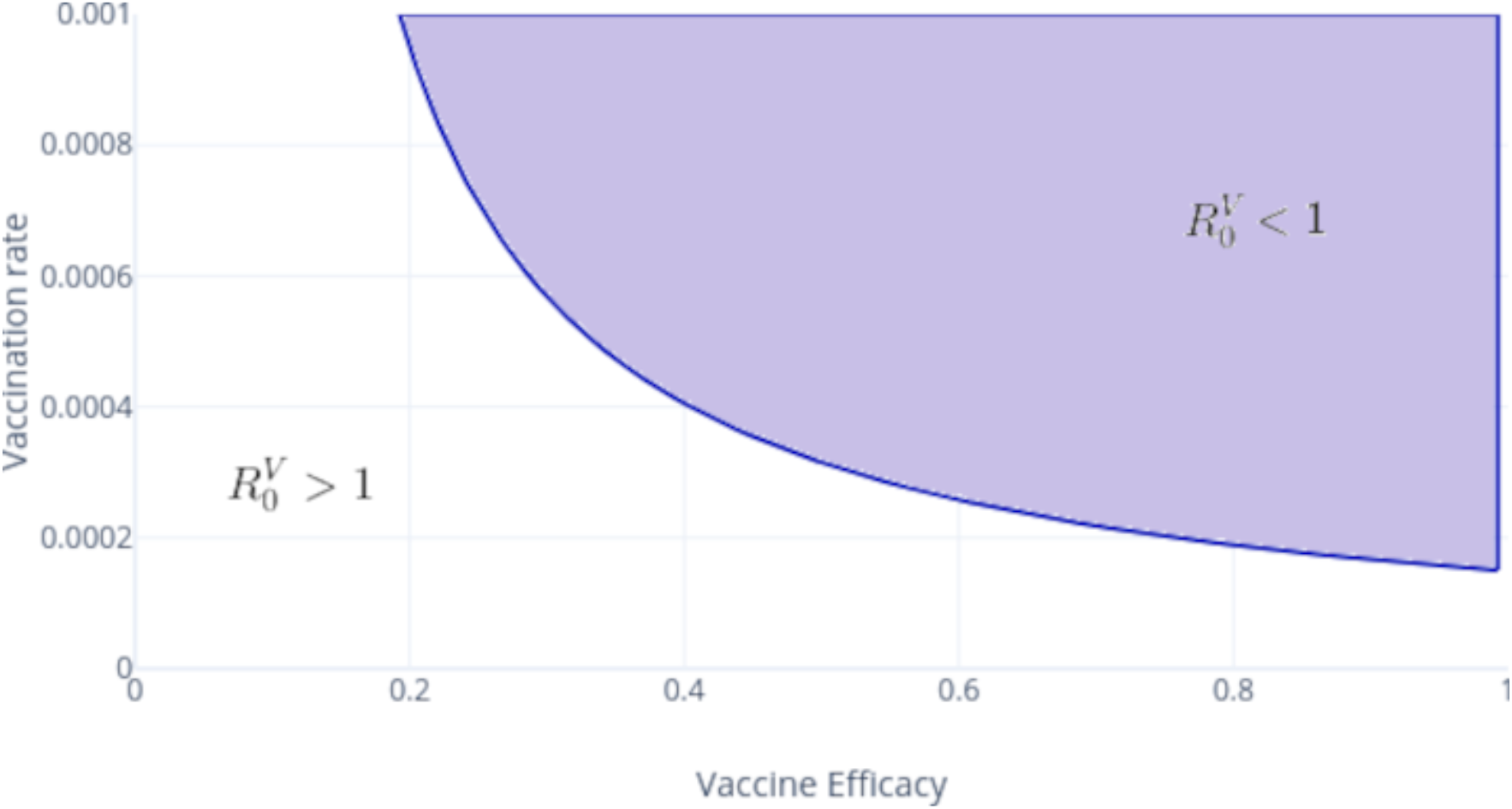
Vaccine efficacy versus vaccination rate feasibility. In the purple shaded region 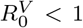 and in the white region 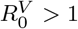. Note that, for this scenario, we consider lockdown individuals. https://plotly.com/AdrianSalcedo/76/

## 5. Optimal controlled version

Now we model vaccination, treatment, and lockdown as an optimal control problem. According to dynamics in Equation (1), we modulate the vaccination rate with a time-dependent control signal *u*_*V*_ (*t*). We add compartment *X*_*vac*_ to count all the vaccine applications of lockdown susceptible, exposed, asymptomatic, and recovered individuals. This process is modeled by

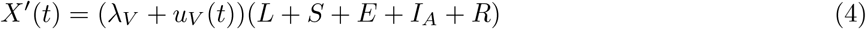

and describes the number of applied vaccines at time *t*. Consider

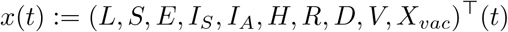

and control signal *u*_*v*_(·). We quantify the cost and reward of a vaccine strategy policy via the penalization functional

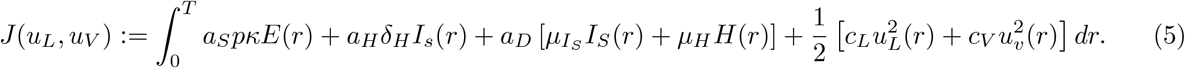

In other words, we assume in functional *J* that pandemic cost is proportional to the symptomatic hospitalized and death reported cases and that a vaccination and lockdown policies implies quadratic consumption of resources.

Further, since we aim to simulate vaccination policies at different coverage scenarios, we impose the vaccination counter state’s final time condition *X*_*vac*_(*T*)

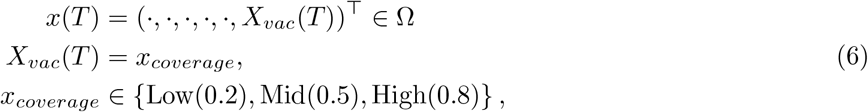

where Ω is the positive invariant set

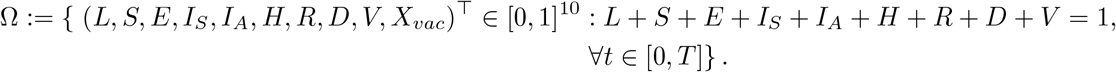

Thus, given the time horizon *T*, we impose that the last fraction of vaccinated populations corresponds to 20%, 50% or 80%, and the rest of final states as free. We also impose the path constraint

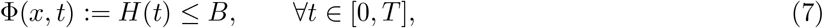

to ensure that healthcare services will not be overloaded. Here *κ* denotes hospitalization rate, and *B* is the load capacity of a health system.

Given a fixed time horizon and vaccine efficiency, we estimate the constant vaccination rate as the solution of

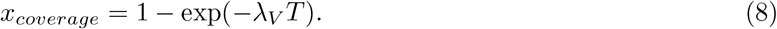

That is, *λ*_*V*_ denotes the constant rate to cover a fraction *x*_*coverage*_ in time horizon *T*. Thus, according to this vaccination rate, we postulate a policy *u*_*v*_ that modulates vaccination rate according to *λ*_*V*_ as a baseline. That is, optimal vaccination amplifies or attenuates the estimated baseline *λ*_*V*_ in a interval 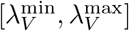 tooptimize functional *J* (·)—minimizing symptomatic, death reported cases and optimizing resources.

Our objective is minimize the cost functional (5)—over an appropriated functional space—subject to the dynamics in equations (1) and (4), boundary conditions, and the path constraint in (7). That is, we look for vaccination policies *u*_*V*_ (·), which solve the following optimal control problem (OCP)

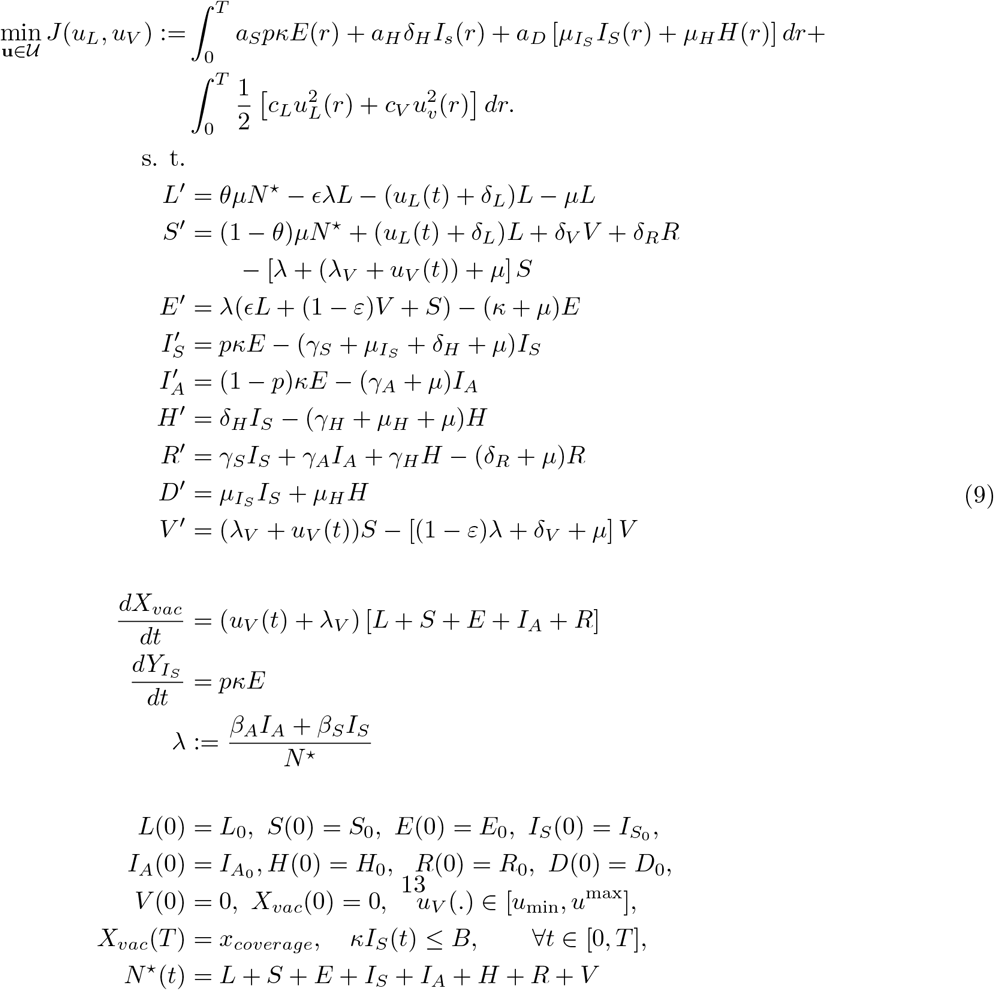

## 6. Numerical Experiments

### 6.1 Methodology

We simulate a scenario corresponding to a hypothetical but plausible initial conditions and parameters. We integrate model in Equation (9) by classic Runge Kutta scheme and solve the optimization stage with the so-called Differntial Evolution method. Differential Evolution (DE) [22] is an evolutionary algorithm successfully employed for global optimization [23]. The method is designed to optimize functions *f*: ℝ*n* → ℝ. Nevertheless, DE can be applied to optimize a functional as stated in [24]. The method can be coded following Algorithm 1, where an initial random population on the search space 𝒱 of size *N*_*p*_ is subjected to mutation, crossover and selection. After this process a new population is created which, again would be subjected to the evolutionary process. This process is repeated until some stopping criteria is fulfilled. Finally the best individual (according to some objective function *f*_*ob*_ to optimize) is extracted. These operations are conducted by the operators **X**_0_, **M, C, S, x**_*best*_; whose explicit form are coded in [25].

In the optimization of this study, the mutation scale factor *F* and the crossover probability *C*_*r*_ were taken as 1 and 0.3 respectively, additional *N*_*p*_ has been taken as 4 times the number of parameters (the dimension of the vector used to describe the two controls—see [24]), which in our case was of 180. As stopping criteria we have used a maximum number of generations which is taken as 5000.

We provide in [26] a GitHub repository with all regarding R and Fortran sources for the sake of reproductivity. This repository also encloses data sources and a Wolfrang Mathematica notebook to reproduce all reported figures.

#### Algorithm 1

Differential Evolution Algorithm

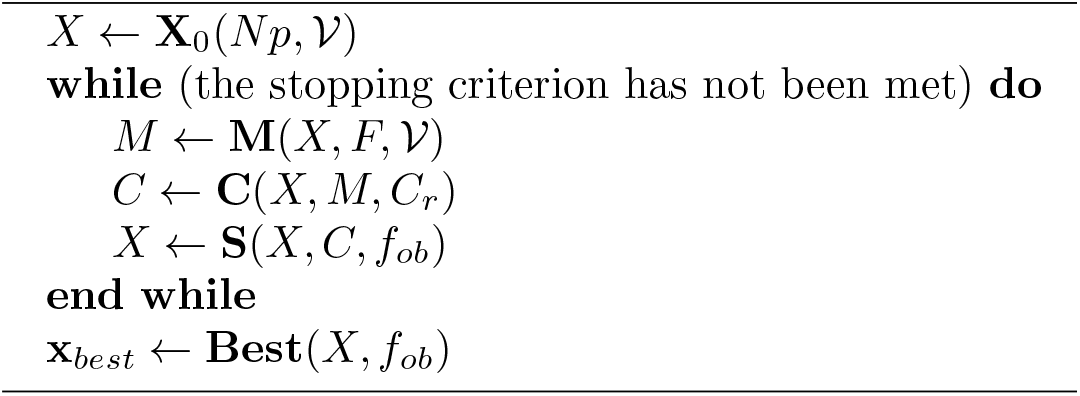

### 6.2 Simulation

According to official Governmental communication in December, Mexico treated 36 000 000 doses Pfizer-Biotech, 76 000 000 doses with Aztra Seneca 18 000 000 quantities of Cansino-BIO. Other developments also are running the third Phase, and with high probability, in the third quarter of 2021, some of these developments will incorporate into Mexico’s vaccine portfolio. Despite official agreements, each vaccine’s delivery schedule is under uncertainty and-or subject to the approval of COFEPRIS.

The first accepted vaccine—Pfizer-BioNTech’s BNT162b2—has an efficacy above 90 % and requires two doses to achieve immunity. The other mentioned developments have a very similar profile but require different logistic management and stock allocation. Thus, we face designing a dose application schedule subject to a given vaccine stock applied in a given period. To this end, we solve the optimal control problem (9). We understand as solution of this problem a pair (*x*(·), *u*(·)) where

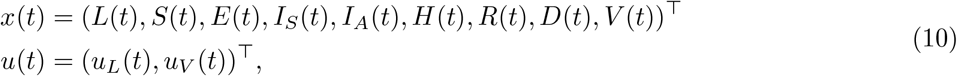

minimizes the burden of COVID-19 in DALYs [9] and the quadratic application cost of the lockdown-vaccination policy formulated in the functional (5).

### Simulation Scenario

#### Initial Conditions

We assume and a hypothetical scenario where the whole population faces a second wave of COVID-19. Thus whole epidemiological classes have positive prevalence. We also assume that Lockdown and Susceptible compartments initially enclose more than 70 % of the total population, and the initial prevalence of symptomatic cases is below critical levels. Further, under our assumptions, the outbreaks’ second wave implies a growth accordingly with an Effective Reproductive Number (ERN) above one—Fig 10 displays a qualitative schematic representation.

**Figure 9:**
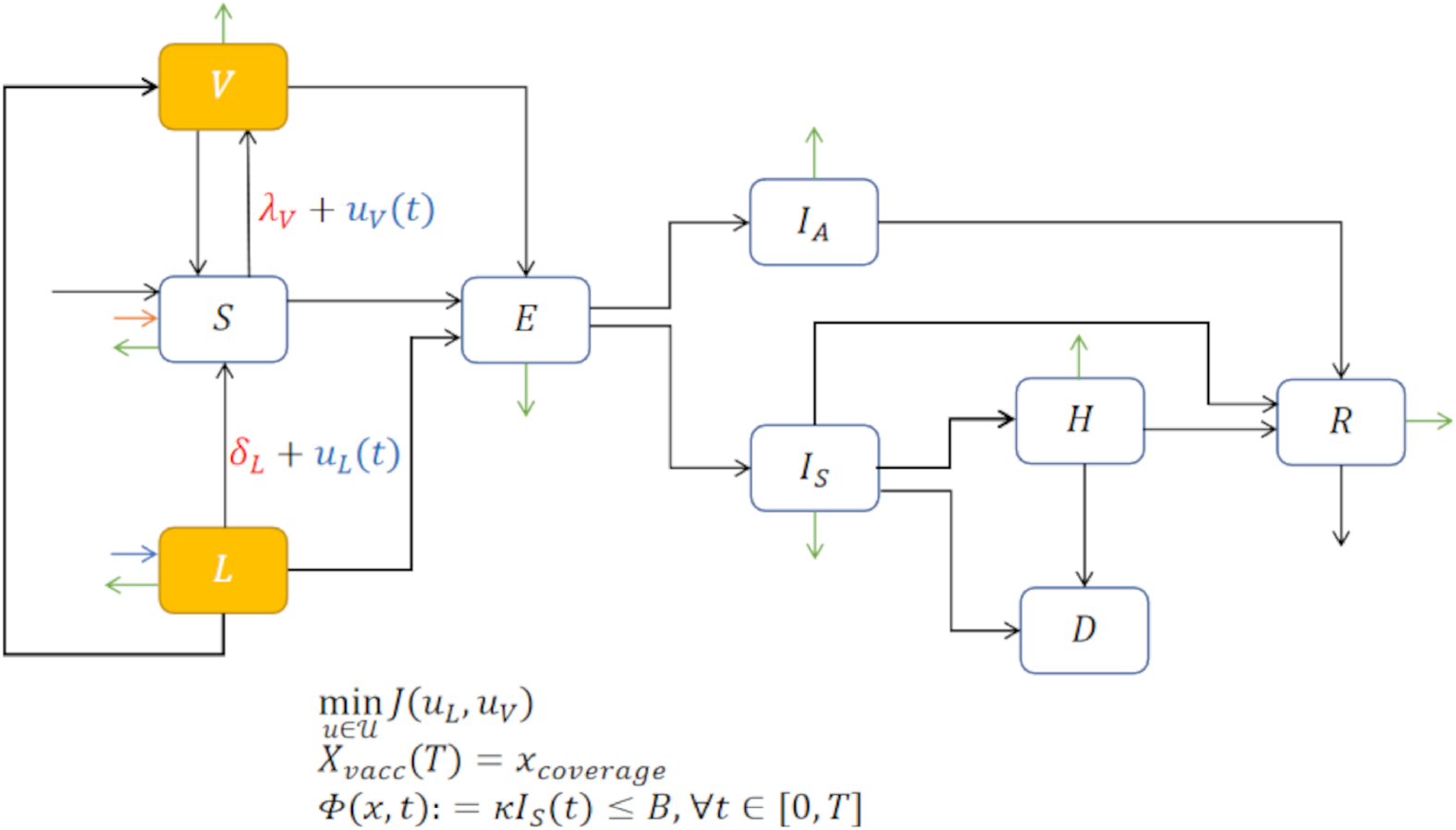
Compartmental diagram of COVID-19 transmission dynamics that includes optimal vaccination dynamics, penalization and a path constraint

**Figure 10:**
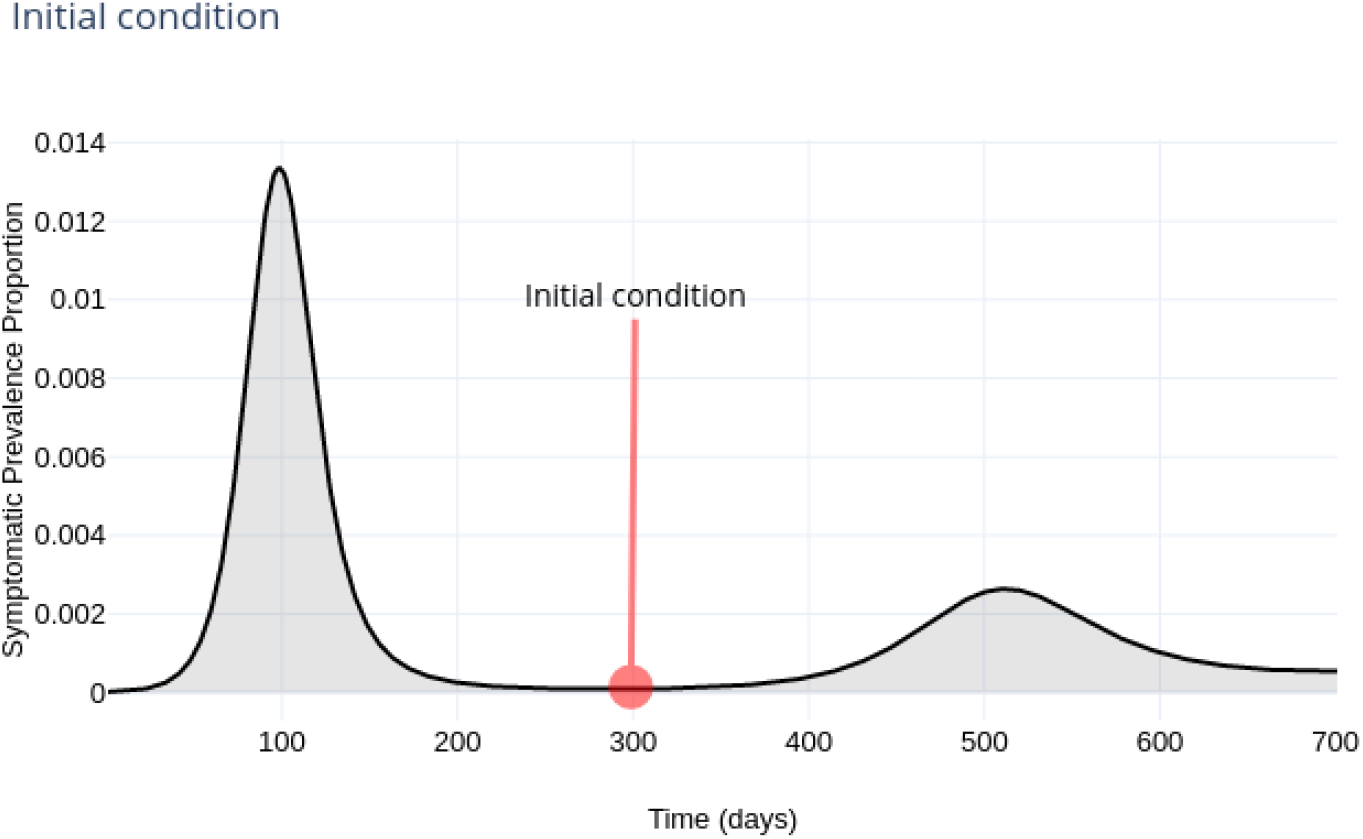
Initial condition scheme. We assume a positive prevalence. For reference, at the date of write this manuscript, prevalence in CDMX is around 16 000 cases, see https://plotly.com/sauld/36/ to display an electronic viewer.

**Figure 11:**
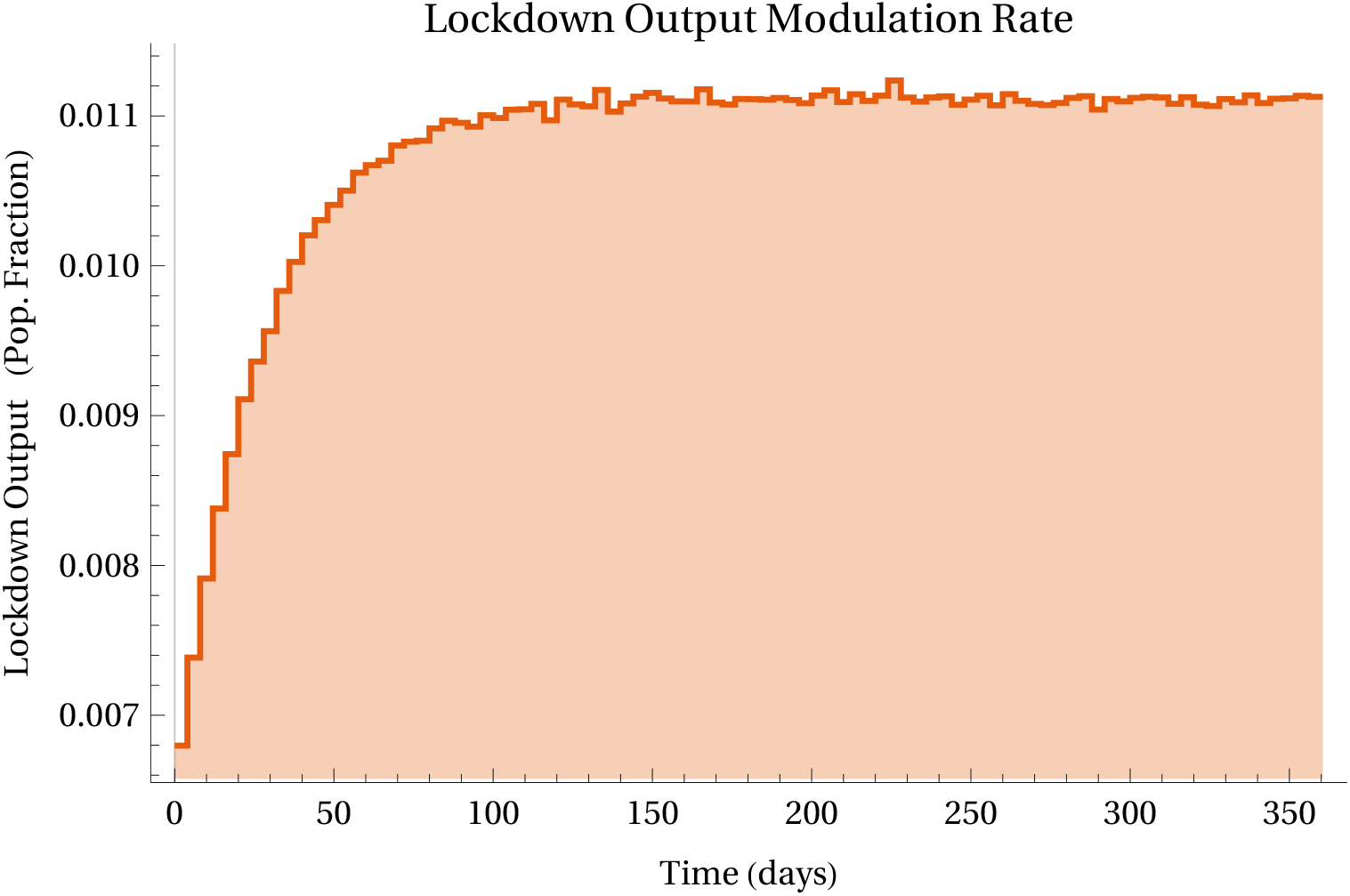
Lockdown modulation signal. https://plotly.com/AdrianSalcedo/56/ to display a electronic viewer.

**Figure 12:**
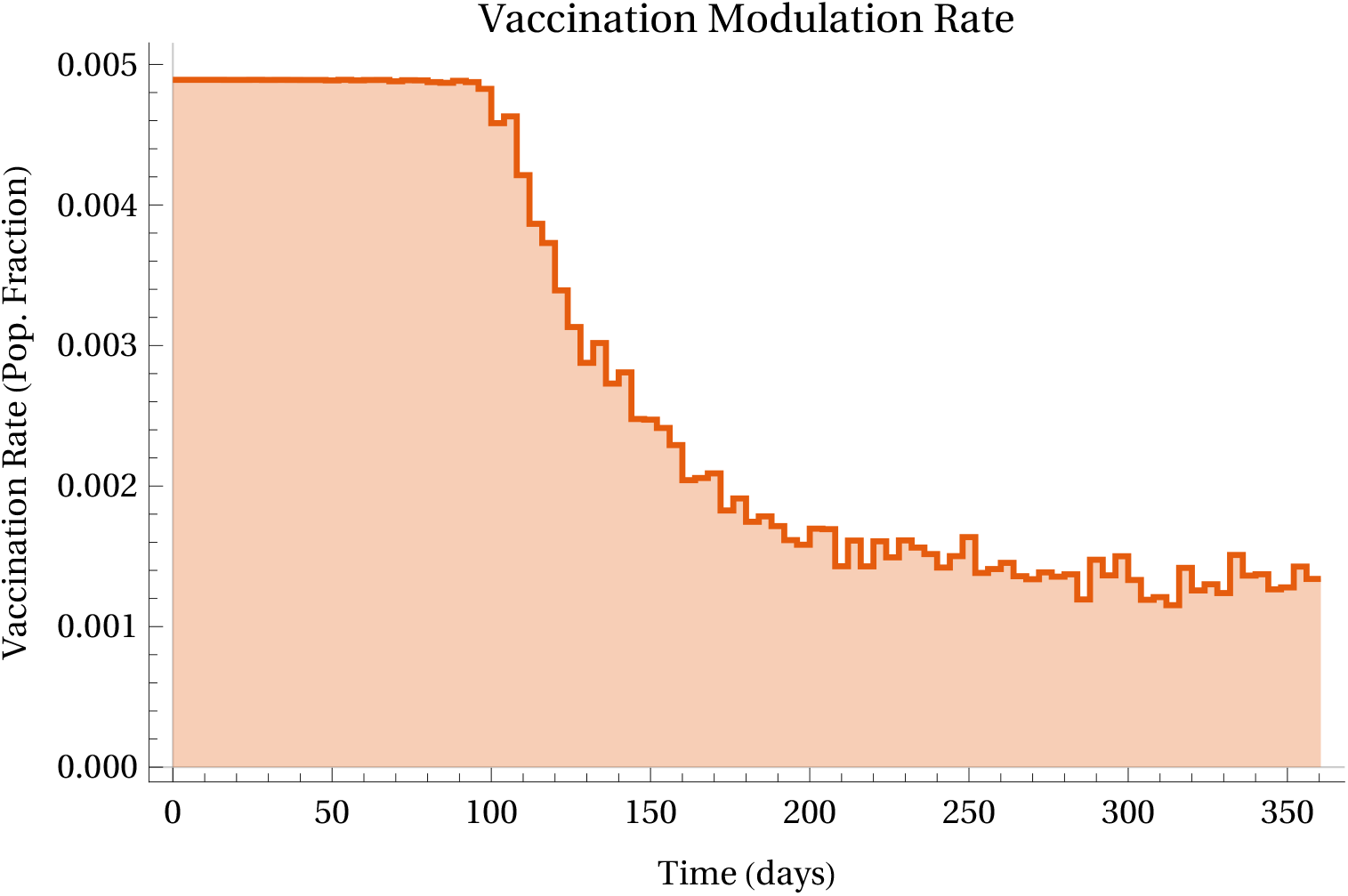
Vaccination rate modulation. https://plotly.com/AdrianSalcedo/58/

**Figure 13:**
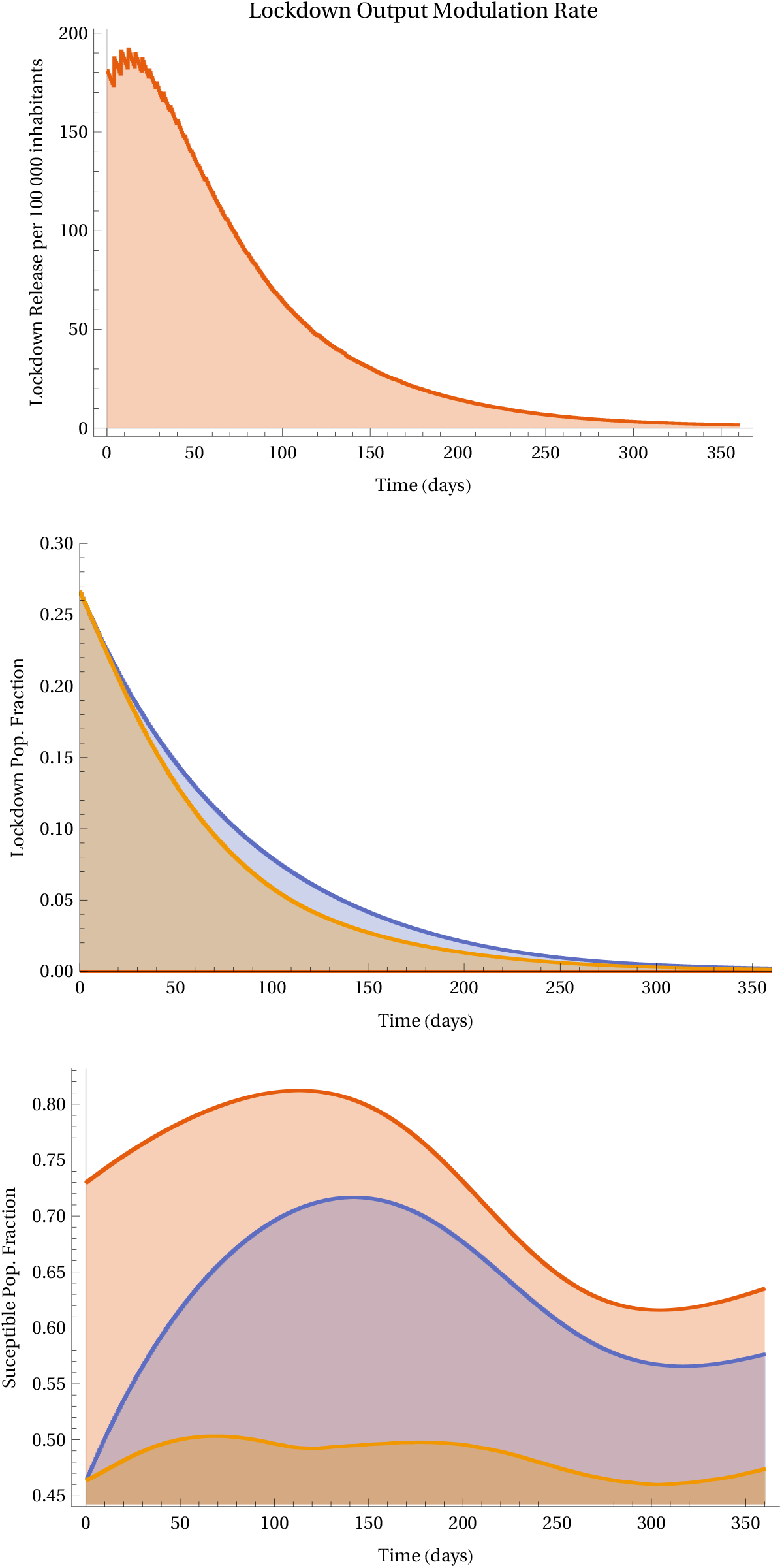
Modulation lock down release. https://plotly.com/AdrianSalcedo/60/

**Figure 14:**
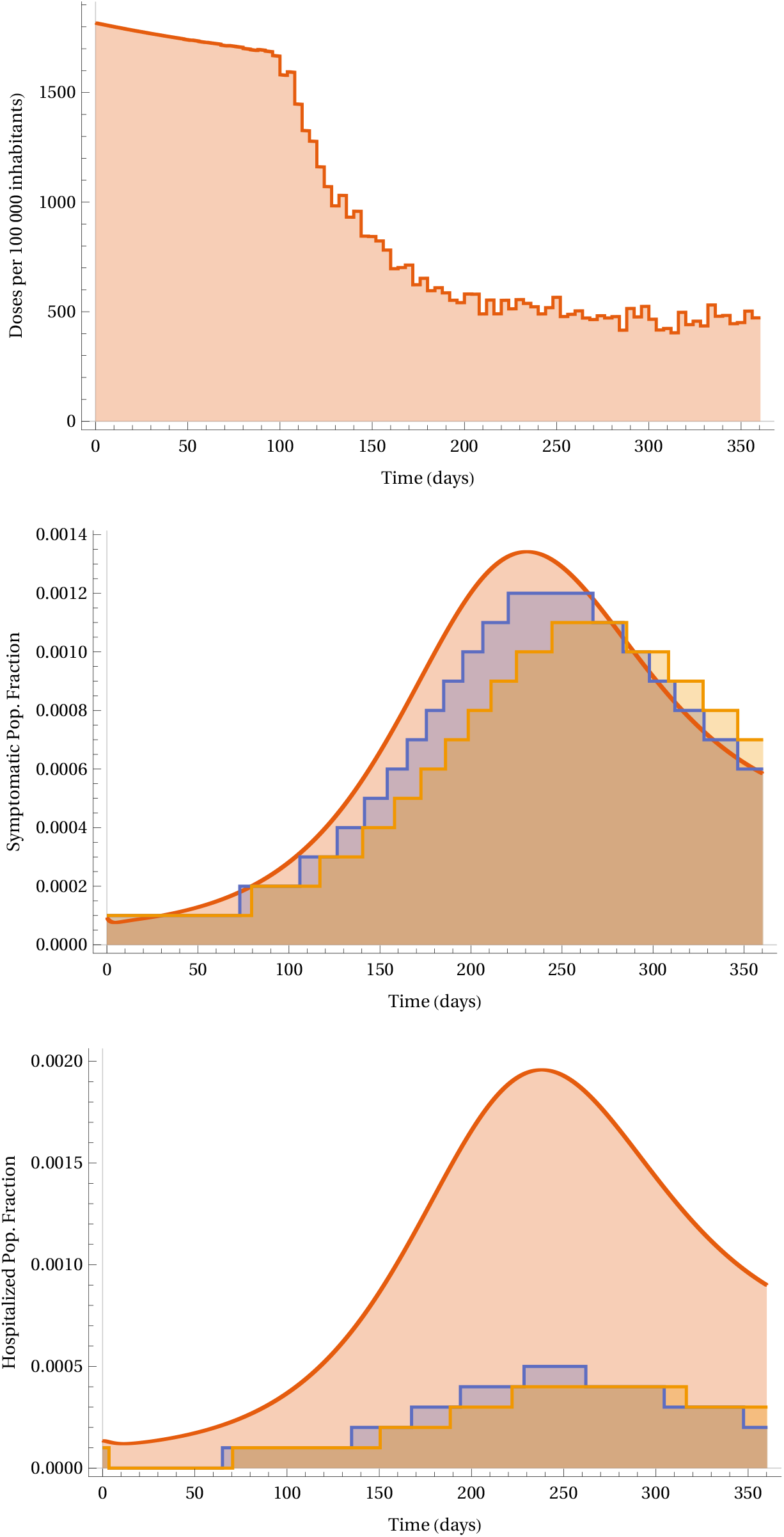
Symptomatic Prevalence and Hozpitalization. https://plotly.com/AdrianSalcedo/61/

## 7. Conclusion

Despite that NPIs have been implemented in most countries to mitigate COVID-19, these strategies cannot develop immunity. Thus, vaccination becomes the primary pharmaceutical measure. However, this vaccine has to be effective and well implemented in global vaccination programs, and each development implies particular issues. Thus new challenges in distribution, stocks, politics, vaccination efforts, among others, emerge. In this work, we have studied the effect of the combined strategy lockdown-vaccination and our result suggest that the NPIs would be essential to face the new emergent problems with the accepted vaccines.

We consider that it is very important to implement stratification by risk and age. Thus our efforts in future research would be directed in combination with the results presented in this manuscript.

## Data Availability

Simulation source code in
https://github.com/SaulDiazInfante/NovelCovid19-OptimalPiecewiseControlModelling

## Data availability

https://github.com/SaulDiazInfante/NovelCovid19-OptimalPiecewiseControlModelling.git

## Authors’ contributions

**Gabriel A. Salcedo-Varela** Conceptualization, Methodology, Software, Validation, Formal analysis, Investigation, Resources, Visualization, Project Administration, Writing–original draft, Writing–review & editing.

**F. Peńuńuri** Methodology, Software, Validation, Formal analysis, Investigation, Data curation, Visualization, Supervision, Writing–original draft, Writing–review & editing.

**David González-Sánchez:** Conceptualization, Methodology, Formal analysis, Writing–original draft, Writing–review & editing.

**Saúl Díaz-Infnte:** Conceptualization, Methodology, Formal analysis, Writing–original draft, Writing–review & editing. Methodology, Software, Validation, Formal analysis, Investigation, Data curation, Visualization, Supervision

## Conflicts of interest

The authors have no competing interests.

## Appendix A. Existence of optimal policies

In this appendix, we show the existence of optimal policies in the class of *piecewise constant policies*. Consider the following cost functional that we want to minimize

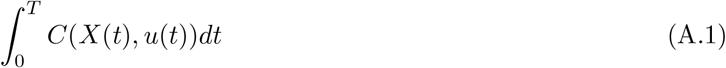

subject to the dynamics

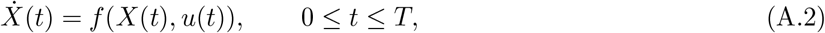

and the initial state *X*(0) = *x*_0_. The functions *u*: [0, *T*] → *U* are called *control polices*, where *U* is a subset of some Euclidean space. Let *t*_0_ *< t*_1_ *< …< t*_*n*_, with *t*_0_ = 0 and *t*_*n*_ = *T*, be a partition of the interval [0, *T*]. We consider piecewise constant policies *ũ* of the form

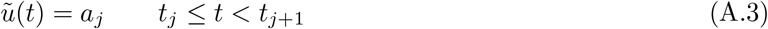

for *j* = 0, *…, n* − 1.

### Assumptions 3.

We made the following assumptions.

(A-1) The function *f* in the dynamics (A.2) is of class *C*^1^.

(A-2) The cost function *C* in (A.1) is continuous and the set *U* is compact.

By Assumption (A-1), the system

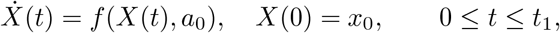

has a unique solution 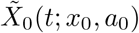 which is continuous in (*x*_0_, *a*_0_); see, for instance [27]. Next, put 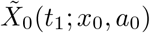 and consider the system

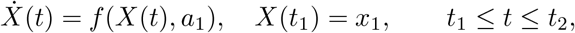

Again, by Assumption (A-1), the latter system has a unique solution 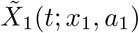 which is continuous in (*x*_1_, *a*_1_). By following this procedure, we end up having a recursive solution

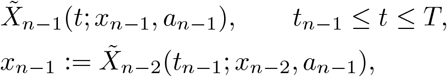

where 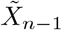 is continuous in (*x*_*n*− 1_, *a*_*n*− 1_).

For a control *ũ* of the form (A.3) and the corresponding solution path 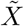, we have

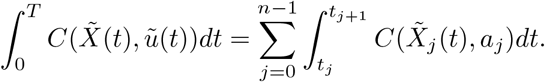

Notice that each 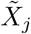 is a continuous function of (*a*_0_, *…, a*_*j*_) and *x*_0_.

By Assumption (A-2), the mapping

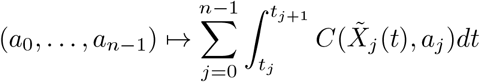

is continuous. Since each piecewise constant policy *ũ* of the form (A.3) can be identified with the vector (*a*_0_, …, *a*_*n*− 1_) in the compact set *U* × … × *U*, the functional (A.1) attains its minimum in the class of piecewise constant policies.

The cost functional (5) and the dynamics (1) are particular cases of (A.1) and (A.2), respectively, and satisfy Assumptions (A-1) and (A-2). Then there exists an optimal vaccination policy of the form (A.3).

## Notes

### Competing Interest Statement

The authors have declared no competing interest.

### Funding Statement

No external funding was received.

### Author Declarations

The authors confirm that all ethical guidelines have been followed. In fact, the nature of this study does not require physical proves.

